# Humoral and T-cell immune response after three doses of mRNA SARS-CoV-2 vaccines in fragile patients: the Italian VAX4FRAIL study

**DOI:** 10.1101/2022.01.12.22269133

**Authors:** Paolo Corradini, Chiara Agrati, Giovanni Apolone, Alberto Mantovani, Diana Giannarelli, Vincenzo Marasco, Veronica Bordoni, Alessandra Sacchi, Giulia Matusali, Carlo Salvarani, Pier Luigi Zinzani, Renato Mantegazza, Fabrizio Tagliavini, Maria Teresa Lupo-Stanghellini, Fabio Ciceri, Silvia Damian, Antonio Uccelli, Daniela Fenoglio, Nicola Silvestris, Fausto Baldanti, Giulia Piaggio, Gennaro Ciliberto, Aldo Morrone, Franco Locatelli, Valentina Sinno, Maria Rescigno, Massimo Costantini, on behalf of the VAX4FRAIL Study Group

## Abstract

**Background:** Patients with solid or hematological tumors, neurological and immune-inflammatory disorders represent potentially fragile subjects with increased risk to experience severe COVID-19 and inadequate response to SARS-CoV2 vaccination.

**Methods:** We designed a prospective Italian multicentric study to assess humoral and T-cell response to SARS-CoV2 vaccination in patients (n=378) with solid tumors (ST), hematological malignancies (HM), neurological (ND) and immuno-rheumatological diseases (ID). The immunogenicity of primary vaccination schedule and of the booster dose were analyzed.

**Results:** Overall, patient seroconversion rate after two doses was 62.1%. A significant lower rate was observed in HM (52.4%) and ID (51.9%) patients compared to ST (95.6%) and ND (70.7%); a lower median level of antibodies was detected in HM and ID versus the others (p<0.0001). A similar rate of patients with a positive SARS-CoV2 T-cell response was observed in all disease groups, with a higher level observed in the ND group. The booster dose improved humoral responses in all disease groups, although with a lower response in HM patients, while the T-cell response increased similarly in all groups. In the multivariable logistic model, the independent predictors for seroconversion were disease subgroups, type of therapies and age. Notably, the ongoing treatment known to affect the immune system was associated with the worst humoral response to vaccination (p<0.0001), but had no effects on the T-cell responses.

**Conclusions:** Immunosuppressive treatment more than disease type *per se* is a risk factor for low humoral response after vaccination. The booster dose can improve both humoral and T-cell response.

**Article’s main point:**

- Lower rate of seroconversion was observed in fragile patients as compared to healthy controls
- The booster dose improves humoral and T-cell response in all fragile patient groups
- Immunosuppressive treatment was associated with the worst humoral response to vaccination, but had no effects on T-cell responses.

## Introduction

In immune-compromised patients, coronavirus disease 2019 (COVID-19) has been associated with an increased risk of hospitalization and death in comparison with the general population[1–4]. Vaccines are the best way to decrease disease severity and control pandemic. mRNA-1273 (Moderna) and BNT162b2 (Pfizer BioNTech) vaccines showed high efficacy in preventing COVID-19 in healthy individuals[5,6]. However, patients diagnosed with solid tumours (ST), hematological malignancies (HM), immune-rheumatological diseases (ID) and neurological disorders (ND) were not included in registration trials. Some studies reported a poor humoral response in patients with malignancies and/or diseases that required immunosuppressive therapies after both natural infection[7] and vaccination[8–16]. Probably this impaired immune response varies according to the degree of immunosuppression, depending on the underlying disease and mostly on its specific treatment. While several data on seroconversion are present, the effectiveness of vaccination on antigen-specific T-cell response as well as the effect of a booster dose in the majority of this fragile population remains largely unknown[17,18]. In addition, several groups have reported a potential time-dependent waning of vaccine-induced immune response[19], in healthy subjects, highlighting the need of a booster dose[20]. Recently, the Italian authorities approved the administration of an additional vaccine dose to improve SARS-CoV-2 protection conferred by immunization in fragile patients, including the four categories evaluated in the present study

Thirteen Italian research hospitals [Istituti di Ricovero e Cura a Carattere Scientifico, (IRCCS)] conducted a prospective study (VAX4FRAIL) aimed at evaluating efficacy and safety of mRNA-based vaccination in patients affected by HM, ST, ND, and ID[21]. Here, we present the results on humoral and T-cell immune responses after complete mRNA-based vaccination and after the third additional dose.

## Methods

### Study design

Between March 2021 and August 2021, 570 patients with a diagnosis of HM, ST, ND or ID, eligible for BNT16b2 or mRNA-1273 vaccination were included in the study. The study, according to the National COVID-19 procedures, was approved by the Italian Regulatory Agency (AIFA) and by the Ethics Committee of IRCCS L. Spallanzani (code 304, 2021). The control group consisted of 180 healthy health-care workers (HCWs) matched for sex and age vaccinated at the IRCCS L. Spallanzani. A written consent was obtained by the study participant.

Inclusion criteria were: diagnosis as above, age ≥ 18 years, SARS-CoV-2 mRNA-based vaccination and life expectancy of at least 12 months at the time of vaccine administration. The main exclusion criterion was the presence of a previous laboratory-confirmed SARS-CoV-2 infection (serology and/or molecular test). Given the disease heterogeneity of the study population, patients were also sub-divided in 4 different subgroups according to the expected immune impairment attributable to their immunosuppressive treatment. Complete description of subgroup categories according to underlying conditions and immunosuppressive treatment are reported in supplementary materials.

### Laboratory Procedures

Anti-Spike SARS-CoV-2 antibodies and T-cell response were monitored at 5 time points (Figure 1): the day of first dose administration (T0), the day of second dose administration (T1); between 5 and 7 weeks after T0 for patients receiving Pfizer/BioNTech vaccine and between 6 and 8 weeks after T0 for patients receiving Moderna vaccine (T2); the day of the booster dose (T pre-3D) and 3 or 4 weeks after (T post-3D).

**Figure 1:**
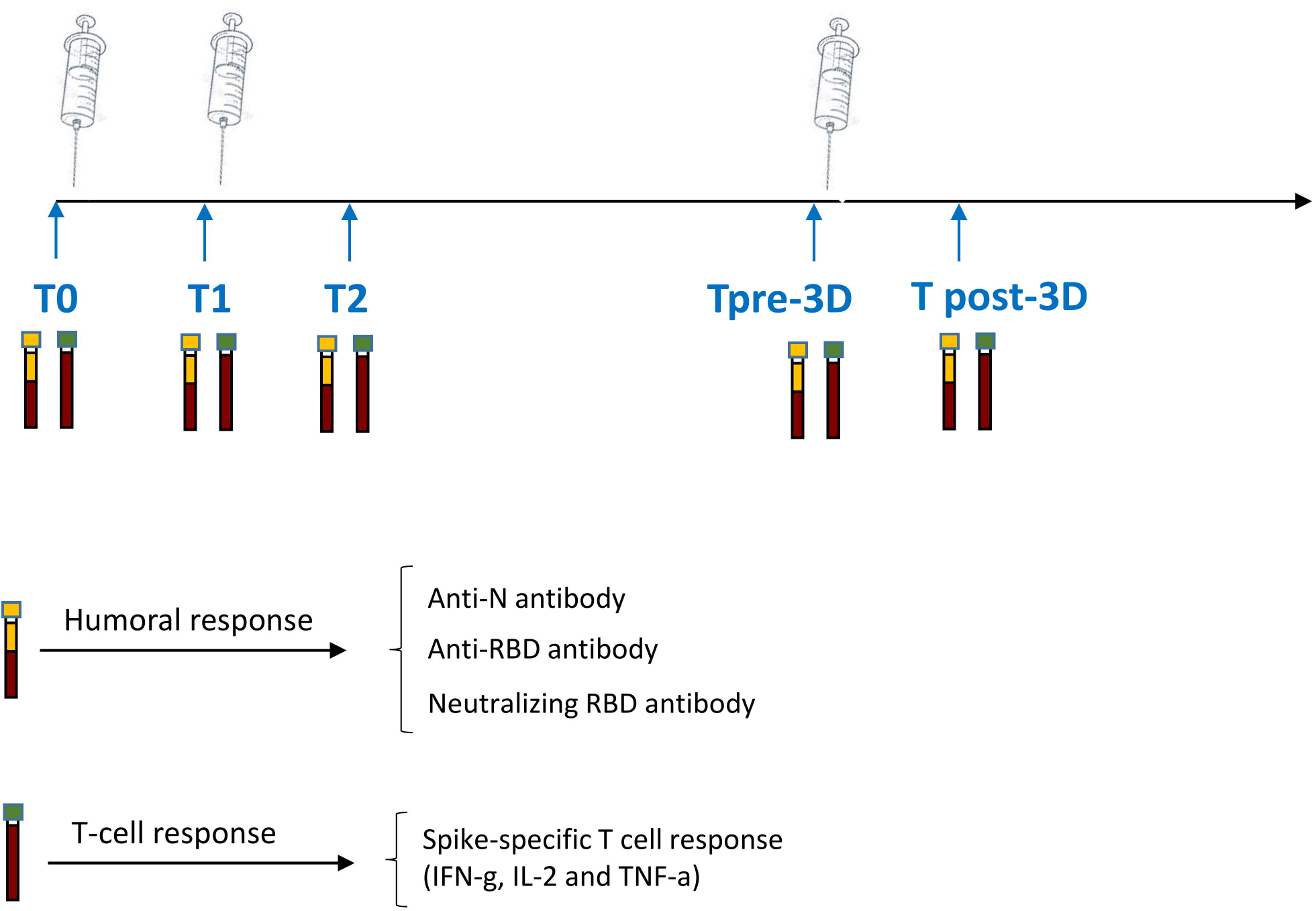
Study design. Schematic representation of the timeline of immune-monitoring of the clinical study

The primary endpoint of the study was to assess the seroconversion rate at T2, compared with the HCW evaluated at the same time point interval after immunization. The secondary endpoints were the evaluation of humoral and cellular responses at each time point in the fragile population and comparing these levels with those achieved by HCWs. The immune response was evaluated also in accordance to disease and treatment specific subgroups, as defined above. Finally, secondary endpoints included evaluation of neutralization activity of vaccine-induced anti-Spike antibodies. The humoral (anti-Nucleoprotein-IgG and the anti-RBD-IgG, neutralizing antibody) and cell mediated immune response was performed as previously described [22] and detailed in the Supplementary materials.

### Statistical Methods

Quantitative variables were summarized using median and Interquartile Range (IQR) while categorical variables were reported using absolute counts and percentage. Differences in seroconversion rate across subgroups were tested using the chi-square and from a multivariable logistic regression model we obtained odds ratio and their 95% confidence intervals.

The Mann-Whitney test was used to assess differences in antibody titers and correlations among humoral and cellular immunity were evaluated through the Spearman’s rho correlation coefficient. IBM SPSS vers.20.0 statistical software was used for analysis.

## Results

### Patient characteristics

Between March-August 2021, 570 patients and 180 HCW were enrolled into the Vax4Frail study; 465 patients received BNT162b2, whereas 105 received mRNA-1273 vaccine.

One hundred and ninety-five of 570 patients were excluded because they did not meet the inclusion criteria or because the samples were not collected at all the pre-specified time points. The analysis of immunogenicity was therefore conducted on a final cohort of 375 patients. The median age of the population was 59 (range 19-86), and 209 participants (55.7%) were female. One hundred patients (26.7%) were affected by HM, 114 (30.4%) had ST, 79 (21.0%) had ID and 82 (21.9%) subjects had ND. A detailed description of patients’ characteristics is available in Table I.

**Table I:**
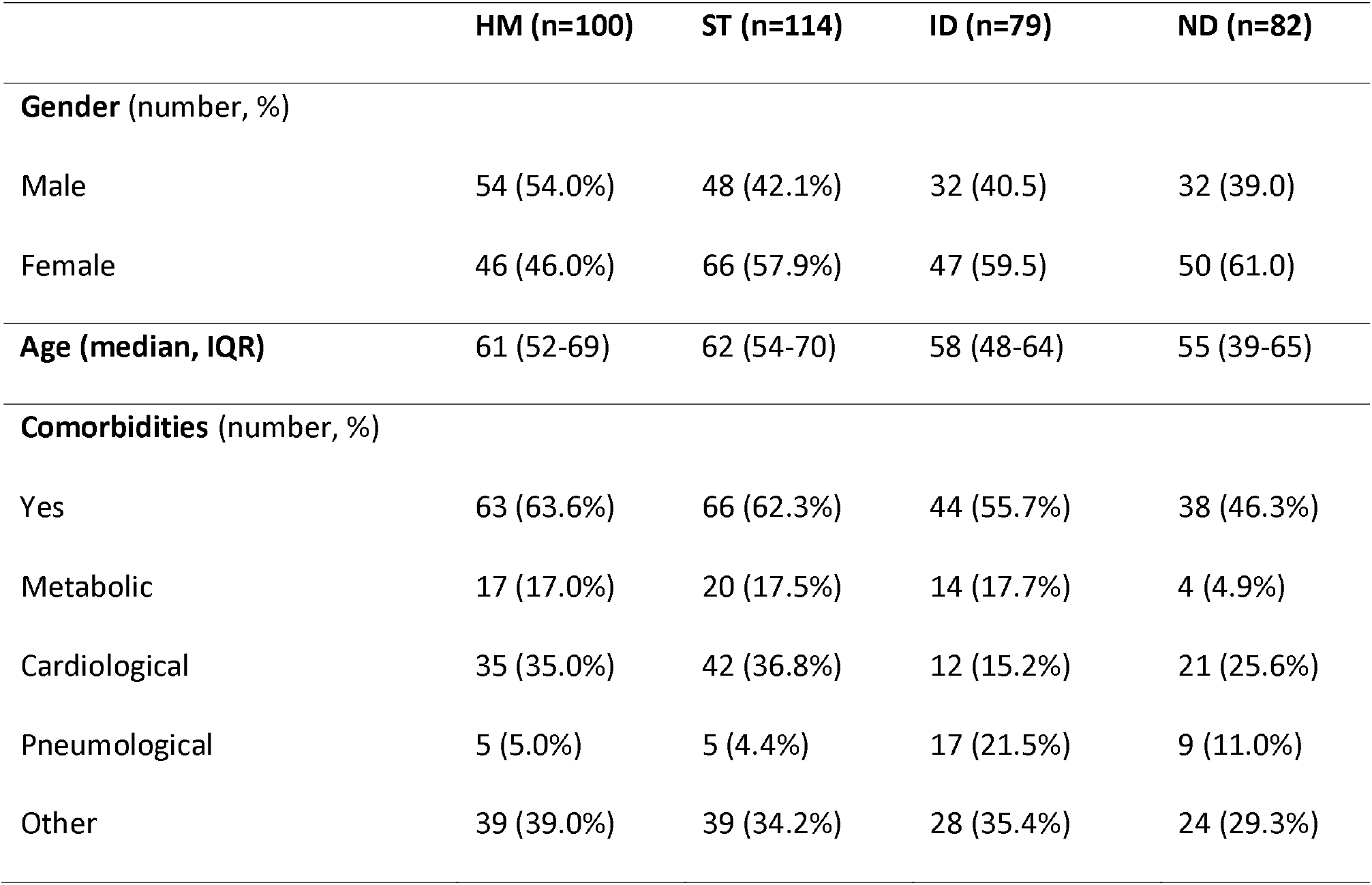
Patients characteristics. The demographic and clinical features of enrolled patients are described. HM: Hematological malignancies; ST: Solid Tumors; ID: Immunorheumatological Diseases; ND: Neurological Disorders

### Impact of different diseases on the humoral response to vaccination

Overall, 259 patients (69.1%, 95% CI 64.4-73.7) seroconverted after the second dose (T2), a significant lower proportion if compared to HCW (100%, p<0.00001). Accordingly, a significant lower median of anti-RBD antibody titer at T2 was observed in patients when compared to HCW [median patients: 172.8 (IQR: 0.7-1387.0) vs median HCW: 2405 (IQR: 1343.0-3848.0), p<0·0001].

The 4 patient groups showed a different kinetic of humoral response, leading to a different frequency of responder patients, as described in Figure 2A. Specifically, patients affected by HM and ID had a significant lower seroconversion rate at T2 (52·4%, CI 42.2-61.8 and 51.9%, 95% CI 39.6-61.6, respectively) compared to ST, and ND patients (95·6%, CI 92.4-98.8 and 70.7%, 71.8% CI 60.9-80.6, respectively, p<0.00001).

**Figure 2:**
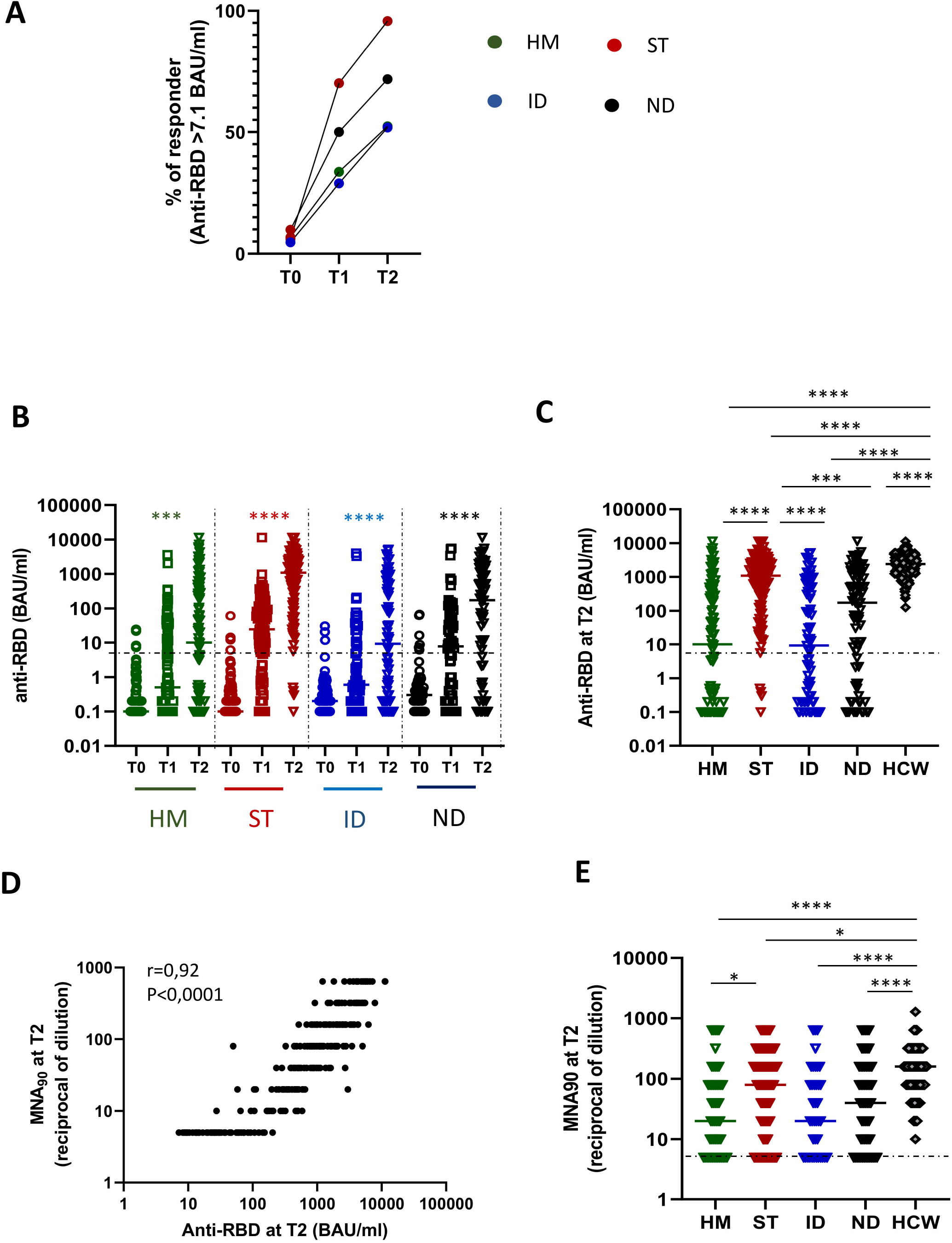
Impact of different diseases on humoral response. **A**: The percentage of patients (HM, green dot; ST, red dots; ID, blue dots and ND, black dots) presenting a positive anti-RBD response (> 7.1 BAU/ml) at each time point (T0, T1 and T2) is shown. **B**: Kinetics of humoral immune response before and after vaccination in HM (green dots), ST (red dots), ID (blue dots) and ND (black dots). SARS-CoV-2 specific anti-RBD Abs were measured in sera samples at each time point. Anti-RBD-IgG are expressed as BAU/ml and values >7.1 BAU/ml are considered positive. Differences were evaluated by Friedman paired test. **** p<0.0001. **C**: The level of anti-RBD antibodies at T2 was compared among groups and was expressed as BAU/ml. Differences were evaluated by Kruskall-Wallis test. *: p<0.05; **: p<0.01; *** p<0.001; **** p<0.0001. HM: median = 10.0 BAU/mL (IQR 0.1-392.5 BAU/mL); ST: 1094.6 BAU/mL (IQR 265.1-2697.9 BAU/mL); ID: 9.2 BAU/mL (IQR 0.2-503.8 BAU/mL); NT: 172.9 BAU/mL (IQR 1.7-1457.8 BAU/mL) and HCW: 2405.0 BAU/ml (IQR 1343-3848 BAU/ml, respectively). **D**: The correlation between the levels of anti-RBD and neutralization titer at T2 for all fragile patients are shown. Each black dot represents one sample. Spearman test: rho = 0.9202, p<0.0001. **E:** The levels of neutralizing antibody at T2 were quantified by microneutralization assay (MNA_90_) in all groups and were expressed as reciprocal of dilution. Differences were evaluated by Kruskall-Wallis test. *: p<0.05; **** p<0.0001. HM: median = 20 reciprocal of dilution (IQR 5-80); ST: 80 reciprocal of dilution (IQR 20-240); ID: 20 reciprocal of dilution (IQR 5-80)); NT: 40 reciprocal of dilution (IQR 8.75-160) and HCW: 160 (IQR 80-320).

In each group, the vaccination was effective in improving the humoral response, inducing a significant increase of anti-RBD antibody (p<0.0001 for each group, Figure 2B). Nevertheless, a lower anti-RBD titer at T2 was observed in HM and in ID when compared to ST, ND and HCW (p<0.001). We therefore measured the neutralizing activity of anti-RBD antibody against SARS-CoV-2 infectivity in BSL-3 facility. This assay was performed on anti-RBD positive samples. Results showed that the percentage of anti-RBD-positive patients showing a neutralizing activity at T2 was 73% (HM), 80,7% (ST), 69,2% (ID) and 74,9% (ND). Of note, a positive correlation between anti-RBD and neutralization titer was observed (rho = 0.92, p<0·0001, Figure 2D). Accordingly, all diseases groups had a significantly lower titer of neutralizing antibody than HCW, and values in HM patients were significantly lower than in ST patients (p<0.0001; Figure 2E).

Detailed humoral response according to different subgroups within HM, ST, IR, and ND groups are reported in Supplementary Tables I-IV. Briefly, among HM patients those treated with B-cell depleting therapies had the lowest seroconversion rate (0%) and the lowest median antibody titre (median = 0.01 BAU/mL, IQR 0.01-0.04). Higher seroconversion rate was reported among all different ST patient subgroups. Considering ID subjects, the lowest vaccine immunogenicity and the lowest antibody levels were detected in patients with ANCA-associated vasculitis or interstitial lung disease on treatment with anti-CD20 monoclonal antibody with or without corticosteroids (25.0%, 95% CI 11.6-38.4 and median = 0.02 BAU/mL, IQR 0.01-0.06). Also in the setting of ND patients, the lowest humoral response rate was documented in individuals exposed to anti-CD20 monoclonal antibody due to multiple sclerosis, with a seroconversion rate of 39.4% (95% CI 22.7-51.1) and median antibody titer of 0.03 U/mL (IQR 0.01-0.10) (Suppl. Tables I-IV).

### Impact of different diseases on the T-cell response to vaccination

A lower frequency of patients showing a positive Spike-specific T-cell response (defined as IFN-γ levels ≥ 12 pg/mL) was detected respect to HCW (80.0%, 95% CI 75.9-84.0 vs 100%, p<0.001). Of note, a positive T-cell response was observed in 218 (84.2%) and 82 (70.7%) patients who did or did not show seroconversion respectively (p=0.003) and only 34 (9.1%) were negative for both types of immune responses.

Subsequently, we assessed the kinetics of the spike-specific T-cell response before and after vaccination in the disease groups (Figure 3A). Results showed that the level of S-specific T-cell response significantly increased overtime in all groups (p<0.0001). Nevertheless, HM, ST and ID showed a significant lower IFN-γ production at T2 when compared to both ND and HCW (p<0.05, Figure 3B). The Spike-specific T-cell response in ND was comparable to HCW. Among fragile patients, IFN-γ values were directly correlated with IL-2 and TNF-α levels (rho= 0.87 and rho = 0.63, p<0.0001 for both), suggesting a coordinated T-cell response to vaccination (Figure 3C). No correlation was observed between anti-RBD and T-cell response (not shown). Detailed T-cell response according to different subgroups are reported in supplementary materials.

**Figure 3:**
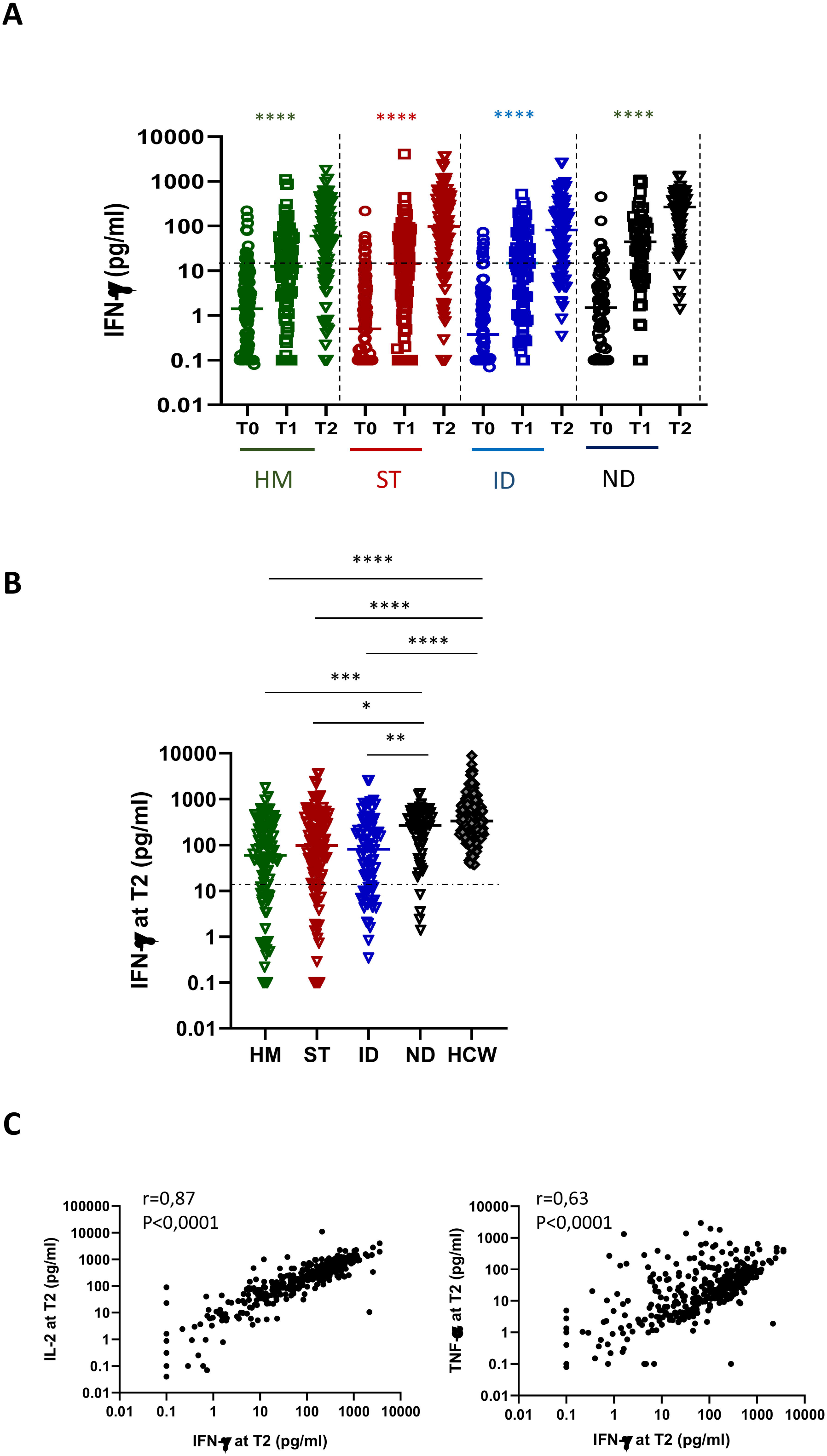
Impact of different diseases on T cell response. **A** Kinetic of T-cell response before and after vaccination in HM (green dots), ST (red dots), ID (blue dots) and ND (black dots). Spike-specific T cell response was measured after stimulation of whole blood with specific peptides at each time point. T cell response was expressed as pg/ml of IFN-γ and values >12 pg/ml are considered positive. Differences were evaluated by Friedman paired test. **** p<0.0001. **B**: The level of T cell response at T2 was compared among groups and was expressed as pg/ml of IFN-γ. Differences were evaluated by Kruskall-Wallis test. *: p<0.05; **: p<0.01; *** p<0.001; **** p<0.0001. HM: median = 60.2 pg/ml (IQR 9.4-247.2 pg/ml); ST: 98.6 pg/mL (IQR 18.9-335.1 pg/mL); ID: 81.8 pg/mL (IQR 12.1-284.1 pg/mL); NT: 268.5 pg/mL (IQR 107.6-505.5 pg/mL) and HCW: 330.5 pg/ml (IQR 187.3-762.7 pg/ml, respectively). **C:** The correlation between the levels of IFN-γ and IL-2 or IFN-γ and TNF-α at T2 for all fragile patients are shown. Each black dot represents one sample. Spearman test: rho = 0.8739 and 0.6368, p<0.0001.

### Effect of the booster dose on both B- and T-cell response

The median interval between second and third dose was 5 months, according to health authority indications. Samples before (T pre-3D) and after (T post-3D) the third dose of vaccine were collected in a cohort of 120 patients (HM: n=19, ST: n=37; ID: n=37 and ND: n=27), who were therefore evaluable for the effect of booster dose on both humoral and T-cell response.

The level of antibodies decreased at 5 months after the second dose of vaccine in all the four groups (p <0.001) except for ID patients which instead showed a weak albeit not significant increase of antibody titers (Figure 4A). This might suggest that ID patients have a delayed induction of a humoral response. The HM had the lowest antibody level and a large fraction of them became seronegative (74.3%). The neutralization test performed on anti-RBD positive samples confirmed also a significant reduction of protective antibodies in all groups (Figure 4B).

**Figure 4:**
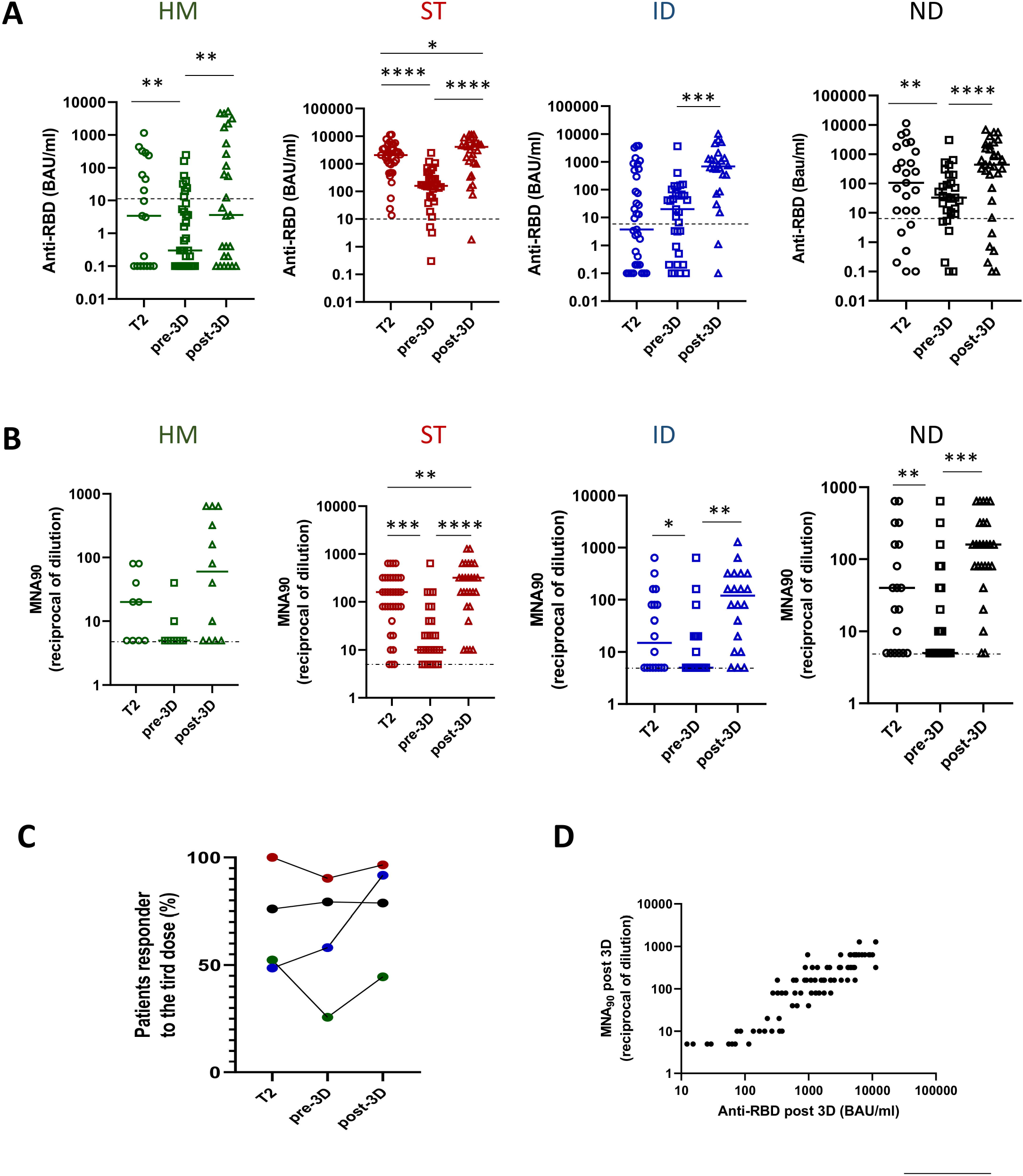
Kinetic of humoral response after two and three doses of vaccine in diseases groups. **A**: The level of the anti-RBD antibody was compared in HM (green dots), ST (red dots), ID (bleu dots) and ND (black dots) at three different time points: after two doses (T2) and before (pre-3D) and after (post-3D) the booster dose. Differences were evaluated by Wilcoxon paired test. HM: median T2: 3.4 BAU/mL (IQR 0.1-238.8 BAU/mL); median pre-3D: 0.3 BAU/mL (IQR 0.1-8.1 BAU/mL); median post-3D: 3.6 BAU/ml (IQR 0.1-555.6 BAU/ml). ** p<0.001. ST: median T2: 2089 BAU/mL (IQR 956.7-3652.0 BAU/mL); median pre-3D: 158.1 BAU/mL (IQR 58.0-444.6 BAU/mL); median post-3D: 4093 BAU/ml (IQR 1051.0-5769.0 BAU/ml). *: p<0.05; **** p<0.0001. ID: median T2: 3.7 BAU/mL (IQR 0.1-400.4 BAU/mL); median pre-3D: 20.0 BAU/mL (IQR 0.5-72.7 BAU/mL); median post-3D: 694.0 BAU/ml (IQR 154.0-1356.0 BAU/ml). *** p<0.001. ND: median T2: 107.0 BAU/mL (IQR 7.8-1510.0 BAU/mL); median pre-3D: 32.6 BAU/mL (IQR 9.3-151.9 BAU/mL); median post-3D: 443.0 BAU/ml (IQR 48.0-1770.0 BAU/ml). ** p<0.01, **** p<0.0001. **B**: The level of the neutralizing antibody was compared in HM (green dots), ST (red dots), ID (bleu dots) and ND (black dots) at three different time points: after two doses (T2) and before (pre-3D) and after (post-3D) the booster dose. Differences were evaluated by Wilcoxon paired test. HM: median T2: 20 reciprocal of dilution (IQR 5-60); median pre-3D: 5 reciprocal of dilution (IQR 5-7.5); median post-3D: 60 reciprocal of dilution (IQR 5-560). ST: median T2: 160 reciprocal of dilution (IQR 80-320); median pre-3D: 10 reciprocal of dilution (IQR 6.2-40.0); median post-3D: 320 reciprocal of dilution (IQR 80-640). ** p<0.01, *** p<0.001, **** p<0.0001. ID: median T2: 15 reciprocal of dilution (IQR 5-100); median pre-3D: 5 reciprocal of dilution (IQR 5-20); median post-3D: 120 reciprocal of dilution (IQR 12.5-320). * p<0.05, ** p<0.01. ND: median T2: 40 reciprocal of dilution (IQR 5-160); median pre-3D: 5 reciprocal of dilution (IQR 5-40); median post-3D: 160 reciprocal of dilution (IQR 80-320). ** p<0.01, *** p<0.001. **C**: The percentage of patients (HM, green dot; ST, red dots; ID, blue dots and ND, black dots) presenting a positive anti-RBD response (> 7.1 BAU/ml) at T2, before (pre-3D) and after (post-3D) the booster dose is shown. **D**: The correlation between the levels of anti-RBD and neutralization titer at T post-3D for all fragile patients is shown. Each black dot represents one sample. Spearman test: rho: 0.8965, p<0.0001).

After the third dose, the prevalence of anti–SARS-CoV-2 antibodies in the entire population showed a slight, but not significant increase from 67·0% (95% CI 57.4-76.7) to 81·5% (95% CI 73·0-89.9). Of note, 33% of patients who failed to respond to the first two doses, showed a seroconversion after the third dose.

Moreover, the third dose was effective in increasing the titer of anti-RBD in all disease groups, although with different strength (Figure 4A). In HM patients the seroconversion rate persisted dramatically low (44.5%), while in ID patients it strongly increased reaching 90% (Figure 4C). Similar results were confirmed when quantifying the neutralization titer (Figure 5B). Specifically, the neutralizing titers after the third dose showed a positive correlation with anti-RBD data (rho = 0.8965, p<0·0001, Figure 4D) and showed a significant improvement in all groups (p<0.1) except for HM (Figure 4B).

**Figure 5:**
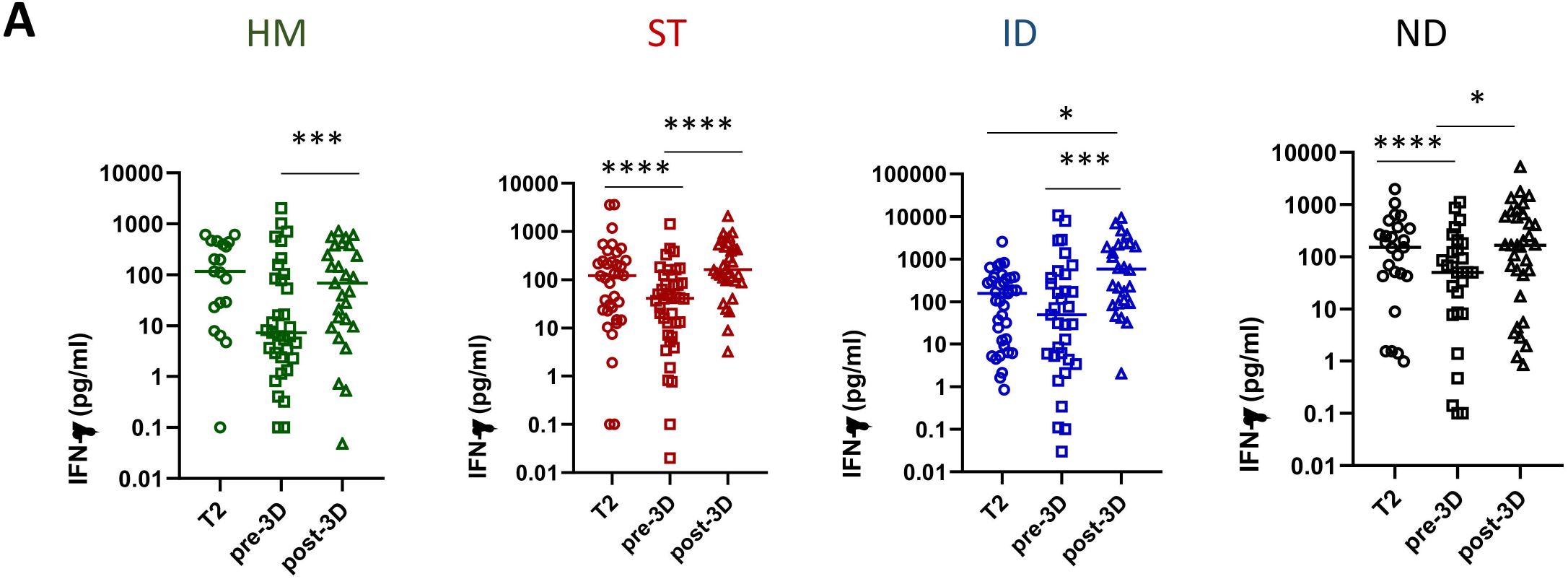
Kinetics of T cell response after two and three doses of vaccine in diseases groups. **A:** S-specific-T cell response expressed as pg/ml of IFN-γ was quantified before and after the booster dose in HM, ST, ID and ND. Differences were evaluated by Wilcoxon paired test. HM: median T2: 116.5 pg/ml (IQR 23.2-436.6 pg/ml); median pre-3D: 7.24 pg/ml (IQR 2.3-94.6 pg/ml); median post-3D: 68.5 pg/ml (IQR 9.5-378.3 pg/ml). *** p<0.001. ST: median T2: 122.8 pg/ml (IQR 23.0-250.8 pg/ml); median pre-3D: 41.3 pg/ml (IQR 10.0-104.7 pg/ml); median post-3D: 163.1 pg/ml (IQR 95.2-522.7 pg/ml). **** p<0.0001. ID: median T2: 158.45 pg/ml (IQR 10.6-354.8 pg/ml); median pre-3D: 49.4 pg/ml (IQR 4.8-402.3 pg/ml); median post-3D: 590.2 pg/ml (IQR 92.4-2223.0 pg/ml). * p<0.05, *** p<0.001. ND: median T2: 152.8 pg/ml (IQR 42.9-358.6 pg/ml); median pre-3D: 50.6 pg/ml (IQR 8.3-170.3 pg/ml); median post-3D: 167.5 pg/ml (IQR 31.6-624.5 pg/ml). * p<0.05, *** p<0.001.

The T-cell response decreased overtime in all groups, but significantly in ST and in ND patients after the first two doses (p<0.01, Figure 5A). The third dose was able to improve T-cell response in all disease groups (p<0.0001), including HM which. In the subgroup of patients receiving the third dose, the percentage of “double negative” subjects decreased from 7.8% to 1.3%.

### Impact of different treatments on the immune response to vaccination

We postulated that treatments damaging the immune system could have an impact on the response to vaccination. Therefore, we decided to divide all patients according to the received/ongoing treatment and its presumed related immunosuppression (Table II).

**Tabel II:** Patients were grouped in 4 different subgroups according to expected immune impairment attributable to their immunosuppressive treatment

|  | **HM (n=100)** | **ST (n=114)** | **ID (n=79)** | **ND (n=82)** |
| --- | --- | --- | --- | --- |
| **No therapy** | 20 (20.0%) | 5 (4.6%) | 0 (0%) | 4 (4.9%) |
| **Low Risk \*** | 30 (30.0%) | 57 (52.3%) | 1 (1.3%) | 24 (29.3%) |
| **Medium Risk<sup>§</sup>** | 15 (15.0%) | 47 (43.1%) | 37 (46.8%) | 21 (25.6%) |
| **High Risk<sup>°</sup>** | 35 (35.0%) | 0 (0%) | 41 (51.9%) | 33 (40.2%) |

\* **Low Risk:** anti-CD30 MoAb, checkpoint inhibitors MoAb; target therapies; hypo-methylation agents, or corticosteroids
<sup>§</sup> **Medium Risk:** chemotherapy (ongoing or in the last 6 months); Bruton's tyrosine kinase inhibitors; BCL-2 inhibitors; anti-CD38 MoAb without immunomodulatory drugs (IMiDs); immunosuppressive agents like methotrexate, mycophenolate mofetil, azathioprine, and cyclosporine.
<sup>°</sup> **High Risk:** Anti-B-cell therapy (ongoing or in the last 12 months): anti-CD20 monoclonal antibody (MoAb) or anti-CD19 chimeric antigen receptor T-cell therapies); allogeneic HSCT

Serological and T-cell response to the first two and to the third dose were then evaluated (Figure 6). Treatments of the high-risk group, i.e. treatments with predicted high damage to the immune response, were associated with the lowest humoral response rate to the first two doses (22.9%, 95% CI 15.0-30.8) when compared to those of the intermediate and low risk groups (84.2%, 95% CI 77.6-90.7, p <0.0001, and 97.3%, 95% CI 94.3-100, p<0.0001, respectively). Similar results were obtained for the response to the third dose: high-risk groups showed the lowest humoral response (41.2%) when compared to those of the intermediate and low-risk groups (90.0%, p=0.0003 and 100%, p<0.0001).

**Figure 6:**
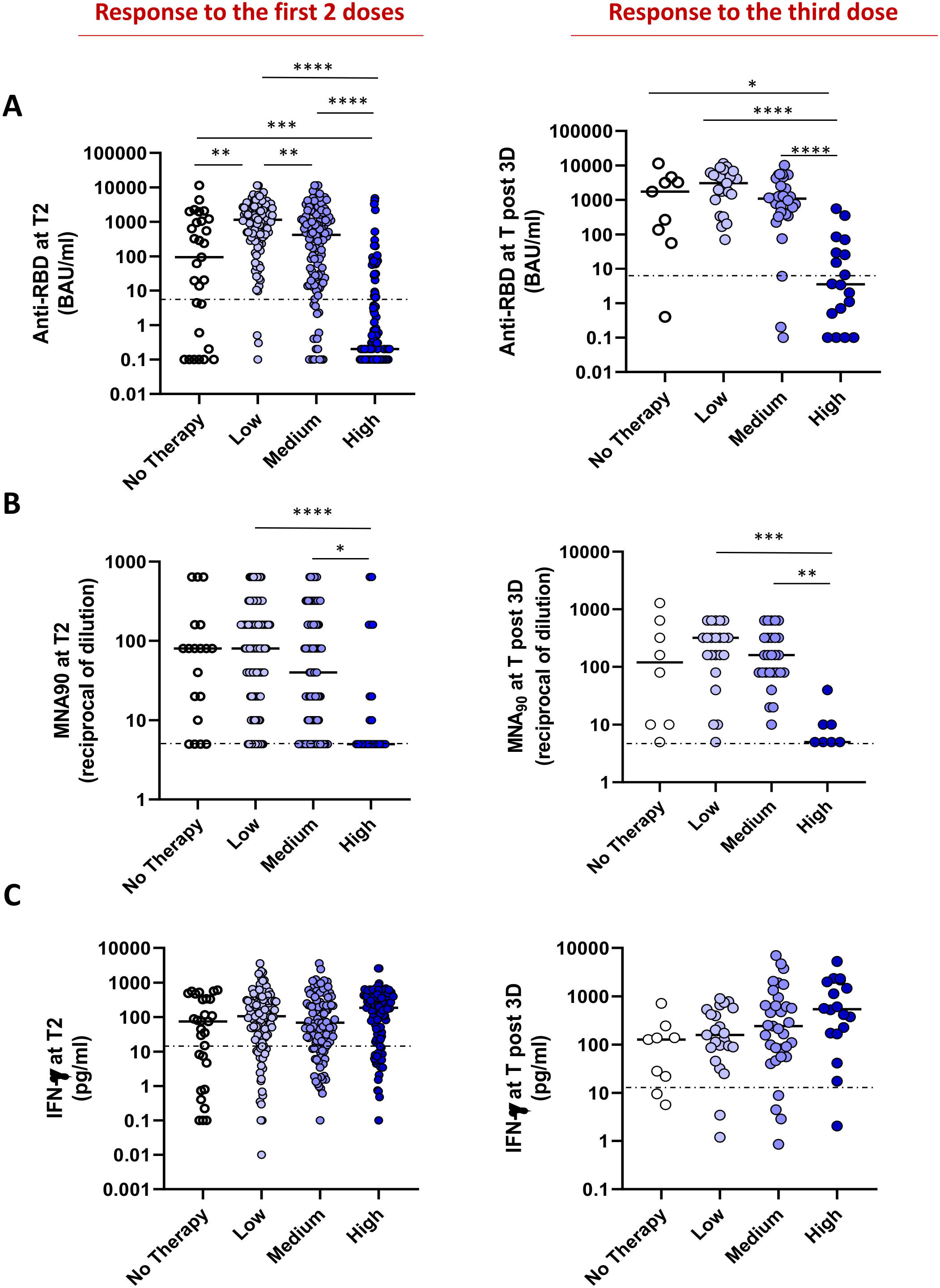
Impact of different therapy on immune response. Independently from the diseases, the patients were divided on the basis of therapy in 4 groups: untreated (white dots), or treated by therapy with a low (light violet dots), medium (dark violet dots) and high (blue dots) impact on the immune system. The immunogenicity of two or three doses of vaccine was compared among the four groups. **A:** The levels of anti-RBD in the four groups are shown. Differences were evaluated by Kuskall-Wallis test. **: p<0.01; *** p<0.001; **** p<0.0001. Response after two doses: No therapy: median = 94.2 BAU/ml (IQR 0.4-1133,0 BAU/ml); Low Impact: 1144.0 BAU/ml (IQR 368.1-11360.0 BAU/ml); Medium Impact: 420.3 BAU/ml (IQR 17.4-1563.0 BAU/ml); High Impact: 0.2 (IQR 0.1-6.4 BAU/ml). Response after the third dose: No therapy: median = 1748 BAU/ml (IQR 95.4-3917.0 BAU/ml); Low Impact: 3044 BAU/ml (IQR 998.8-6175.0 BAU/ml); Medium Impact: 1088.0 BAU/ml (IQR 380.1-2536.0 BAU/ml); High Impact: 3.5 (IQR 0.4-39.6 BAU/ml). **B:** The level of neutralizing antibodies in the four groups is shown. Differences were evaluated by Kruskall-Wallis test. *: p<0.05; **** p<0.0001. Response after two doses: No therapy: median = 80 reciprocal of dilution (IQR 10-160); Low Impact: 80 reciprocal of dilution (IQR 20-160); Medium Impact: 40 reciprocal of dilution (IQR 6.2-160.0); High Impact: 5 reciprocal of dilution (IQR 5-20). Response after the third dose: No therapy: median = 120 reciprocal of dilution (IQR 10-560); Low Impact: 320 reciprocal of dilution (IQR 140-400); Medium Impact: 160 reciprocal of dilution (IQR 80-320); High Impact: 5 reciprocal of dilution (IQR 5-10). **C:** The T cell response, analyzed by quantifying IFN-γ in the four groups is shown. Differences were evaluated by Kruskall-Wallis test. Response after two doses: No therapy: median = 74.9 pg/ml (IQR 2.7-338.4 pg/ml); Low Impact: 105.9 pg/ml (IQR 26.3-291.9 pg/ml); Medium Impact: 69.2 pg/ml (IQR 18.1-297.3 pg/ml); High Impact: 186.3 pg/ml (IQR 22.7-390.0). Response after the third dose: No therapy: median = 127.2 pg/ml (IQR 15.8-192.0 pg/ml); Low Impact: 158.9 pg/ml (IQR 87.2-536.4 pg/ml); Medium Impact: 243.4 pg/ml (IQR 69.3-799.7 pg/ml); High Impact: 545.3 pg/ml (IQR 171.9-2049.0).

Accordingly, a lower anti-RBD titer to the first two and the booster dose was observed in patients treated with high impact therapy when compared with the other groups (p<0.0001, Figure 6A). The neutralization assay, performed on anti-RBD positive patients, showed a lower activity in high-risk group, highlighting a heavily dampened humoral response, which also failed to neutralize the spike protein (Figure 6B). In agreement, in the multivariable logistic model (Table III), the independent predictors for seroconversion were individual subgroups, different therapies and age.

**Table III:** Multivariable logistic model at T2.

|  | **OR (95% CI)** | **P value** |
| --- | --- | --- |
| **Gender** |  |  |
| Male | 0.97 (0.51-1.84) | 0.93 |
| Female | 1 |  |
| **Age (in years)** | **0.97 (0.95-0.99)** | **0.026** |
| **Comorbidities** |  |  |
| Yes | 0.90 (0.45-1.78) | 0.76 |
| No | 1 |  |
| **Subgroups** |  |  |
| Haematological | 1 |  |
| Solid Tumors | 8.09 (2.38-27.53) | **0.001** |
| Immunorheumatological | 2.36 (0.94-5.91) | 0.068 |
| Neurological | 3.72 (1.45-9.54) | **0.006** |
| **Current therapy (impact on immune system)** |  |  |
| No therapy | 1 |  |
| Low | 16.75 (3.85-72.85) | **<0.0001** |
| Medium | 1.98 (0.65-6.07) | 0.23 |
| High | 0.112 (0.04-0.35) | **<0.0001** |

In striking contrast, the T-cell response was similar in all groups, regardless of treatment (Figure 6C).

## Discussion

In this prospective multicenter trial, we found a suboptimal immune response induced by BNT16b2 or mRNA-1273 vaccines. Moreover, this study provides a detailed evaluation of both humoral and cellular response after the booster dose in a large cohort of fragile patients. After the first 2 doses, the overall seroconversion rate was significantly lower compared to the one of HCW. Among the 4 disease groups, HM and ID patients showed the lowest prevalence of anti-SARS-CoV-2 antibodies, with response rates similar to the ones reported by other studies[8,9,23]. This finding is largely due to the detrimental effect of anti B-cell therapies on humoral response. This result highlights the greater impact of treatment rather than the disease itself. In fact, treatment-defined subgroups were more capable than disease-defined ones to predict humoral response after vaccination. The negative effect of B-cell depleting therapies, resulting mostly from anti-CD20 MoAbs, lasted up to 12 months after the end of treatment and this is in line with other reports[17,24,25]. This can be explained by the known prolonged half-life of these biological drugs which are detectable in the serum at lympholytic levels for up to six months after therapy completion and by the subsequent long-lasting B-cell depletion[26,27]. In line with other studies, we reported a high seroconversion rate among ST patients and this is probably due to the use of treatments with lower lympholytic capacity [13,28]. However, even if ST patients had a humoral response, still the antibody titers were lower than those of HCW, suggesting a partially reduced immune response also in this category. Although the precise definition of all the factors responsible for protection against COVID-19 remains to be fully determined, the relationship between *in vitro* neutralization levels and protection from severe COVID-19 has been widely described[29]. Interestingly, we not only reported reduced antibody levels, but also reduced neutralizing antibody levels among all disease subgroups. In contrast to humoral response, the efficacy of the T-cell response in preventing morbidity and mortality of COVID-19 has been less investigated[30], but the role of T-lymphocyte reaction in severe disease has been emphasised by several studies[31]. A recent Italian prospective study evaluated the cellular response in 99 hematologic patients after 2 doses of m-RNA vaccines. A specific T-cell response was detected in 86% and 100% of patients and healthy subjects, respectively. Of note, 74% of seronegative patients had a T-cell response, but both cellular and humoral responses were absent in 13·1% of cases[17]. Our study confirms the rate of T-cell mediated response after 2 doses of vaccine. In addition, we were able to demonstrate the lack of association between humoral and cellular responses and a substantial stability of T-cell response independently from the disease treatment.

Scientific community concurs to the need of a booster dose, given the rapid spread of the variants of concern delta and omicron in addition to the waning immunity to the primary vaccination[32,33]. The greatest benefit from a booster dose is expected to be seen among immunocompromised patients. In particular, some recent studies reported an improved humoral response among immunocompromised subjects[20,34–36]. At 4 weeks after the booster dose, we were able to show an increase of humoral response and neutralizing antibodies. Nevertheless, the seroconversion rate and antibody titers after the third dose were lower in HM than in other diseases, highlighting the peculiar impairment of adaptive immunity in these patients. Of note, differently from HM, ID patients showed an excellent response to the third dose reaching over than 90% of seroconversion rate and an anti-RBD titer higher than that observed after the first two doses. This finding is peculiar and highlights the clinical relevance of the booster dose in this population. ID patients also showed an increase of antibody levels over time after the two doses rather than a decrease, suggesting that this population requires more time to reach a strong B-cell response and this can be improved by a booster dose.

A significant increase of T-cell immune response after the third dose was observed in all disease groups, including HM. The rate of double negative patients dropped to 1.3%, suggesting the development of some potentially protective immune responses in almost all subjects. We suggest that in this small proportion of double negative patients, the prophylactic administration of neutralising anti-spike monoclonal antibodies with prolonged half-life and effectiveness on current and emerging variants might concur to confer protection against SARS-CoV-2.

A limitation of our study is the lack of measurement of neutralization titers against the emerging Omicron variant. However, in this context it has to be taken into account that in healthy subjects a significant increase in the neutralizing response against this variant has been observed after the third dose[37,38].

In conclusion, we demonstrated a lower prevalence of seroconversion among immunosuppressed patients compared to HCW. The lowest humoral response was reported among patients treated with anti-B-cell therapies. However, T-cell response results showed more encouraging data, suggesting a possible benefit of the vaccination due to cellular immunity, particularly in light of the observation that T-cell epitopes are shared among wild type and Omicron variant[39]. Finally, data on the third dose indicate a potential immunological benefit and highlights HM as the major fragile group.

## Data Availability

All data produced in the present study are available upon reasonable request to the authors

## Authors’ contribution

GA, AM and MC conceived the study; CA, PC, CS, PLZ, RM, FT, FC, AU, NS, GC, AM and FL designed the study; PC, MTLS, GP, FB, SD, NS managed the patients; DG performed the statistical analysis; VS monitored the study; CA, VB, AS, GM, VM performed the immunological analysis; CA, MR, DF, PC, FB analysed the immunological data; PC, CA and MR drafted the manuscript; all the authors reviewed and agreed on the final version.

## Acknowledgements

We are indebted for their precious support to this study the Research Director Dr. Giuseppe Ippolito, and the Deputy General Director for Health Research and Innovation Dr. Gaetano Guglielmi, from the Italian Ministry of Health. We would also like to thank the patients who will be enrolled in the VAX4FRAIL study and their families, and all those who will be actively involved in their continuous care, study data collection and analysis, and ultimately in the scientific production that will result from this research.

## THE VAX4FRAIL STUDY GROUP

### PRINCIPAL INVESTIGATORS (alphabetical order)

Giovanni Apolone (Fondazione IRCCS Istituto Nazionale dei Tumori di Milano); Alberto Mantovani (IRCCS Istituto Clinico Humanitas, Milano).

### SCIENTIFIC COORDINATOR

Massimo Costantini (Fondazione IRCCS Istituto Nazionale dei Tumori di Milano).

### STEERING COMMITTEE (alphabetical order)

Chiara Agrati (IRCCS Istituto per le Malattie Infettive Lazzaro Spallanzani, Roma); Giovanni Apolone (Fondazione IRCCS Istituto Nazionale dei Tumori di Milano); Fabio Ciceri (IRCCS Ospedale San Raffaele, Milano); Gennaro Ciliberto (IRCCS Istituto Nazionale Tumori Regina Elena, Roma); Massimo Costantini (Fondazione IRCCS Istituto Nazionale dei Tumori di Milano); Franco Locatelli (Università La Sapienza, Roma); Alberto Mantovani (IRCCS Istituto Clinico Humanitas, Milano); Fausto Baldanti (Fondazione IRCCS Policlinico San Matteo di Pavia); Aldo Morrone (Istituto Dermatologico San Gallicano IRCCS, Roma); Carlo Salvarani (Azienda USL-IRCCS Reggio Emilia); Nicola Silvestris (IRCCS Istituto Tumori “Giovanni Paolo II”, Bari); Fabrizio Tagliavini (Fondazione IRCCS Istituto Neurologico Carlo Besta, Milano); Antonio Uccelli (Ospedale Policlinico San Martino IRCCS, Genova); Pier Luigi Zinzani (IRCCS Azienda Ospedaliero-Universitaria di Bologna).

### DISEASE GROUPS

1. HAEMATOLOGICAL MALIGNANCIES Referent: Paolo Corradini (Fondazione IRCCS Istituto Nazionale dei Tumori, Milano);
2. SOLID TUMORS Referent: Gennaro Ciliberto (IRCCS Istituto Nazionale Tumori Regina Elena, Roma);
3. IMMUNORHEUMATOLOGICAL DISEASES Referent: Carlo Salvarani (Azienda USL IRCCS Reggio Emilia);
4. NEUROLOGICAL DISEASES: Referent: Antonio Uccelli (Ospedale Policlinico San Martino IRCCS, Genova); Renato Mantegazza (Fondazione I.R.C.C.S Istituto Neurologico Carlo Besta (INCB), Milano).

### IMMUNOLOGICAL GROUP

REFERENTS: Chiara Agrati (IRCCS Istituto per le Malattie Infettive Lazzaro Spallanzani, Roma); Maria Rescigno (IRCCS Istituto Clinico Humanitas, Milano); Daniela Fenoglio (Ospedale Policlinico San Martino IRCCS, Genova);

PARTICIPANTS: Roberta Mortarini (Fondazione IRCCS Istituto Nazionale dei Tumori di Milano); Cristina Tresoldi (IRCCS Ospedale San Raffaele, Milano); Laura Conti (IRCCS Istituto Nazionale

Tumori Regina Elena, Roma); Stefania Croci (Azienda USL IRCCS Reggio Emilia); Fausto Baldanti (Fondazione IRCCS Policlinico San Matteo di Pavia); Vito Garrisi (IRCCS Istituto Tumori “Giovanni Paolo II”, Bari); Fulvio Baggi (Fondazione IRCCS Istituto Neurologico Carlo Besta, Milano); Francesca Bonifazi (IRCCS Azienda Ospedaliero-Universitaria di Bologna); Fulvia Pimpinelli (Istituto Dermatologico San Gallicano IRCCS, Roma); Concetta Quintarelli (IRCCS Ospedale Pediatrico Bambino Gesù, Roma); Rita Carsetti (IRCCS Ospedale Pediatrico Bambino Gesù, Roma).

### INMI CENTRALIZED LABORATORY (INMI Lazzaro Spallanzani – IRCCS, Roma) (alphabetical order)

Enrico Girardi (Scientific Director), Aurora Bettini; Veronica Bordoni; Concetta Castilletti; Eleonora Cimini; Rita Casetti; Francesca Colavita; Flavia Cristofanelli; Massimo Francalancia; Simona Gili; Delia Goletti; Giulia Gramigna; Germana Grassi; Daniele Lapa; Sara Leone; Davide Mariotti; Giulia Matusali; Silvia Meschi; Stefania Notari; Enzo Puro; Marika Rubino; Alessandra Sacchi; Eleonora Tartaglia

### CLINICAL TASK FORCE

Paolo Corradini, Silvia Damian, Filippo de Braud (Fondazione IRCCS Istituto Nazionale dei Tumori di Milano); Maria Teresa Lupo Stanghellini, Lorenzo Dagna, Francesca Ogliari, Massimo Filippi (IRCCS Ospedale San Raffaele: Milano); Andrea Mengarelli, Francesco Marchesi, Giancarlo Paoletti e Gabriele Minuti, Elena Papa (IRCCS Istituto Nazionale Tumori Regina Elena, Roma); Elena Azzolini, Chiara Pozzi, Luca Germagnoli, Carlo Selmi, Maria De Santis, Carmelo Carlo-Stella, Alexia Bertuzzi, Francesca Motta, Angela Ceribelli (IRCCS Istituto Clinico Humanitas, Milano); Fausto Baldanti, Sara Monti, Carlomaurizio Montecucco (Fondazione IRCCS Policlinico San Matteo di Pavia); Aldo Morrone (Istituto Dermatologico San Gallicano IRCCS, Roma); Maria Grazia Catanoso, Monica Guberti, Carmine Pinto, Francesco Merli, Franco Valzania (Azienda USL-IRCCS Reggio Emilia); Rosa Divella, Antonio Tufaro, Vito Garrisi, Sabina Delcuratolo, Mariana Miano (IRCCS Istituto Tumori “Giovanni Paolo II”, Bari; Carlo Antozzi, Silvia Bonanno Rita Frangiamore, Lorenzo Maggi (Fondazione IRCCS Istituto Neurologico Carlo Besta, Milano); Antonio Uccelli, Paolo Pronzato, Matilde Inglese, Carlo Genova, Caterina Lapucci, Alice Laroni, Ilaria Poirè (Ospedale Policlinico San Martino IRCCS, Genova); Marco Fusconi, Vittorio Stefoni, Maria Abbondanza Pantaleo (IRCCS Azienda Ospedaliero-Universitaria di Bologna).

### STATISTICAL COMMITTEE

Diana Giannarelli (IRCCS Istituto Nazionale Tumori Regina Elena, Roma).

### e-CRF AND MONITORING REFERENT

Valentina Sinno, Serena Di Cosimo (Fondazione IRCCS Istituto Nazionale dei Tumori di Milano).

### PROJECT MANAGERS OF THE STUDY

REFERENTS: Elena Turola, Azienda USL-IRCCS di Reggio Emilia.

PARTICIPANTS: Iolanda Pulice, Roberta Mennitto Fondazione IRCCS Istituto Nazionale dei Tumori, Milano); Stefania Trinca (IRCCS Ospedale San Raffaele, Milano); Giulia Piaggio (IRCCS Istituto Nazionale Tumori Regina Elena, Roma); Chiara Pozzi (IRCCS Istituto Clinico Humanitas, Milano); Irene Cassaniti (Fondazione IRCCS Policlinico San Matteo, Pavia); Alessandro Barberini (Istituto Dermatologico San Gallicano IRCCS, Roma); Arianna Belvedere (Azienda USL-IRCCS Reggio Emilia); Sabina Del Curatolo (IRCCS Istituto Tumori “Giovanni Paolo II”, Bari); Rinaldi Elena, Federica Bortone (Fondazione IRCCS Istituto Neurologico Carlo Besta, Milano); Maria Giovanna Dal Bello (Ospedale Policlinico San Martino IRCCS, Genova); Silvia Corazza (IRCCS Azienda Ospedaliero-Universitaria, Bologna).

