## Supplementary Materials for "Humoral and T-cell immune response after three doses of mRNA SARS-CoV-2 vaccines in fragile patients: the Italian VAX4FRAIL study"

**Fragile Patients Sub-groups**

Any subject with one of the below reported condition and undergoing SARS-CoV-2 vaccination could be enrolled in the VAX4FRAIL study.

The first cohort of fragile patients consisted of subjects affected by hematological malignancies (HM), due to the low immune response expected secondary to the lymphotoxic treatment required and to the disease itself. These patients were further stratified into 4 groups: i) newly diagnosed requiring treatment; ii) undergoing treatments, iii) treatments completed within 6 months without antibodies (chemotherapy alone, ibrutinib or ruxolitinib) or with antibodies (anti-B-cell therapy, including CAR-T cells, anti-CD30 and PD1 treatments); and iv) patients at least 3 months after autologous or allogeneic transplantation (HSCT) with active immune suppressive therapy.

Also individuals with solid tumour (ST) often have immune deficiencies because of their disease and/or immunosuppressive therapies. These subjects were subdivided in different subgroups: i) individuals treated with adjuvant chemotherapy, ii) chemotherapy in metastatic disease, iii) immunotherapy in metastatic disease and iv) target therapies in metastatic disease.

The ID cohort included patients with history of ANCA-associated vasculitis and interstitial lung disease secondary to connective tissue diseases (systemic sclerosis, mixed connective tissue disease, systemic lupus erythematosus, and Sjogren syndrome), dermatomyositis and polymyositis, and rheumatoid arthritis, treated with traditional immunosuppresant agents or anti-CD20 monoclonal antibody with or without corticosteroids, usually in low doses.

Finally, the ND group consisted of individuals affected by multiple sclerosis treated with anti-CD20 monoclonal antibodies and patients with generalized myasthenia gravis on immunosuppressive therapy (including corticosteroids or B-cell targeted biological treatments).

Given the disease heterogeneity of the study population, patients were also sorted in 4 different subgroups according to the expected immune impairment attributable to their immunosuppressive treatment. Subjects who were on treatment or had received in the 12 months prior to vaccination an anti-B-cell therapy [including anti-CD20 monoclonal antibody (MoAb) or anti-CD19 chimeric antigen receptor T-cell therapies], and those who had received allogeneic HSCT were defined as at high risk of poor response to vaccine; patients who were on treatment or had received chemotherapy in the 6 months prior to vaccination were defined as at intermediate risk. In this group were also included patients treated with Bruton’s tyrosine kinase inhibitors, BCL-2 inhibitors, anti-CD38 MoAb without immunomodulatory drugs (IMIDs), and immunosuppressive therapies like methotrexate, mycophenolate mofetil, azathioprine, and cyclosporine. Subjects who were on treatment with other drugs (e.g anti-CD30 MoAb, checkpoint inhibitors MoAb, target therapies, hypometilating agents, or corticosteroids) were defined as at low risk. In addition, we considered separately patients at diagnosis, who had not yet received any treatment, to assess the impact of the disease on the immune system impairment.

**Immune response to vaccination**

The response to vaccination was assessed by quantifying the anti-Nucleoprotein-IgG and the anti-RBD-IgG (Architect® i2000sr Abbott Diagnostics, Chicago, IL). Anti-N IgG were expressed as a ratio (S/CO) and values were considered positive when ≥ 1.4. Anti-RBD-IgG were expressed as binding arbitrary units (BAU)/mL and values were considered positive when ≥ 7.1. Two- to four-weeks after the second dose as well as before and after the third dose of vaccine, a neutralization assay was performed on anti-RBD positive samples to evaluate the functional activity of vaccination-induced anti-Spike antibodies.

The cell-mediated immune response to vaccination was assessed through a standardized whole blood assay , shared between clinical centers. The Spike specific T cell response was performed by a cytokine-releasing assay after specific stimulation. A concentration equal or more than 12 pg/mL of IFN-γ was considered as positive. Peripheral blood was collected in heparin tubes and stimulated with a pool of peptides spanning the Spike protein (Miltenyi Biotech, Germany) at 37°C (5% CO2). A pool of S protein peptides was generated by combining the PepTivator® SARS-CoV-2 Prot_S, the PepTivator® SARS-CoV-2 Prot_S1 and the PepTivator® SARS-CoV-2 Prot_S+ (all by Miltenyi Biotech) and was used for cell stimulation at the final concentration of 0.1 ug/ml. The spontaneous cytokines release was calculated in unstimulated culture (background) and a superantigen (SEB) was used as positive control. Plasma was harvested after 16-20 h of stimulation and stored at -80°C. Th1-cytokines (IFN-γ, TNF-α, IL-2) were quantified in the plasma samples using an automatic ELISA (ELLA, Protein Simple). The detection limits of these assays were 0.17 pg/ml, 0.3 pg/ml and 0.54 pg/ml for IFN-γ, TNF-α and IL-2, respectively. The results are expressed as the amount of each cytokines after subtracting the background. The cut-off value for each cytokine was defined as the mean +/- 2 SEM of 50 anti-S and anti-N negative HCW before vaccination. A concentration equal or more than 12 pg/mL of IFN-γ was considered as positive. For IL-2 and TNF-α, the cut-off for positivity was identified as 25 pg/mL and 50 pg/mL, respectively.
