## Supplementary Tables for "Humoral and T-cell immune response after three doses of mRNA SARS-CoV-2 vaccines in fragile patients: the Italian VAX4FRAIL study"

**Supplementary Table I: Immune response to vaccination in HM subgroups**

|  | **Responders**  **(n/N, %)** | **Anti-RBD**  **(median, IQR)** | **Double negative**  **(n, %)** | | **Anti IFN-γ**  **responders (n, %)** |
| --- | --- | --- | --- | --- | --- |
| **Subgroup 1 - Hematological malignancies** | 52/100 (52.0%) | 10.0 (0.1-392.5) | 16 (16.0) | 74 (74.0) | |
| **Subgroup 1a - Newly diagnosed patients with ANY haematologic malignancy requiring treatment** | 20/24 (83.3%) | 227.1 (23.9-722.8) | 3 (12.5) | 18 (75.0) | |
| **Subgroup 1b - Pt with ongoing or completed chemotherapy** | 21/26 (80.8%) | 55.2 (9.6-498.8) | 2 (7.7) | 19 (73.1) | |
| **Subgroup 1b - Pt with ongoing or completed Ibrutinib** | 2/6 (33.3%) | 0.4 (0.1-19.4) | 1 (16.7) | 5 (83.3) | |
| **Subgroup 1b - Pt with ongoing or completed ruxolitinib** | 1/2 (50.0%) | 576.6 (0.4-1150.9) | 1 (50.0) | 1 (50.0) | |
| **Subgroup 1c - Pt treated anti-B-cell** | 0/31 (0%) | 0.01 (0.01-0.15) | 8 (25.8) | 23 (74.2) | |
| **Subgroup 1c - Pt treated anti-CD30** | 1/1 (100.0%) |  | 0 | 1 (100.0) | |
| **Subgroup 1c - Pt treated anti-PD1** | 0/0 |  | 0 | 0 | |
| **Subgroup 1d - After allogeneic transplantation** | 2/3 (66.7%) | 197.4 (98.9-200.7) | 1 (33.3) | 0 | |
| **Subgroup 1d - After autologous transplantation** | 5/7 (71.4%) | 2168.4 (140.8-3790.6) | 0 | 7 (100.0) | |

**Supplementary Table II: Immune response to vaccination in ST subgroups**

|  | **Responders**  **(n/N, %)** | **Anti-RBD**  **(median, IQR)** | **Double negative**  **(n, %)** | **Anti IFN-γ responders (n, %)** |
| --- | --- | --- | --- | --- |
| **Subgroup 2 - Solid tumors** | **109/114 (95.6%)** | 1094.6 (265.1-2697.9) | 5 (4.4) | 92 (80.7) |
| **Subgroup 2a - Chemotherapy in adjuvant therapy** | 19/19 (100.0%) | 607.9 (129.2-1358.6) | 0 | 17 (89.5) |
| **Subgroup 2b - Chemotherapy in metastatic disease** | 34/37 (91.9%) | 843.4 (83.2-3472.1) | 3 (8.1) | 28 (75.7) |
| **Subgroup 2c - Immunotherapy in metastatic disease** | 21/22 (95.5%) | 1146.9 (283.9-2536.7) | 1 (4.5) | 17 (77.3) |
| **Subgroup 2d - Target therapies in metastatic disease** | 35/36 (97.2%) | 1570.5 (533.1-26.50.5) | 1 (2.8) | 30 (83.3) |

**Supplementary Table III: Immune response to vaccination in ID subgroups**

|  | **Responders**  **(n/N, %)** | **Anti-RBD**  **(median, IQR)** | **Double negative**  **(n, %)** | **Anti IFN-γ responders (n, %)** |
| --- | --- | --- | --- | --- |
| **Subgroup 3 - Immunorheumatological disease** | **40/79 (50.6%)** | 9.2 (0.2-503.8) | 11 (13.9) | 61 (77.2) |
| **Subgroup 3a - ANCA - associated vasculitis treated with immunodepressive agents with/without glucocrticoids** | 12/15 (80.0%) | 503.8 (14.7-1178.2) | 2 (13.3) | 11 (73.3) |
| **Subgroup 3a - ANCA - associated vasculitis treated with RTX with/without glucocorticoids** | 3/15 (20.0%) | 0.2 (0.1-3.8) | 2 (13.3) | 12 (80.0) |
| **Subgroup 3b - Interstitial lung disease treated with rituximab with/without glucocorticoids** | 7/25 (28.0%) | 0.6 (0.2-20.3) | 3 (12.0) | 20 (80.0) |
| **Subgroup 3b - Interstitial lung disease treated with traditional immunodepressive agents with/without glucocorticoids** | 18/24 (75.0%) | 339.2 (7.7-940.2) | 4 (16.7) | 18 (75.0) |

**Supplementary Table IV: Immune response to vaccination in ND subgroups**

|  | **Responders**  **(n/N, %)** | **Anti-RBD**  **(median, IQR)** | **Double negative**  **(n, %)** | **Anti IFN-γ responders (n, %)** |
| --- | --- | --- | --- | --- |
| **Subgroup 4 - Neurological diseases** | **58/82 (70.7%)** | 172.9 (1.7-1457.8) | 2 (2.4) | 73 (89.0) |
| **Subgroup 4a - Multiple sclerosis with relapsing-remitting MS on Ocrelizumab** | 8/21 (38.1%) | 0.7 (0.1-61.7) | 1 (4.8) | 19 (90.5) |
| **Subgroup 4a - Multiple sclerosis with secondary/primary progressive MS on Ocrelizumab** | 5/12 (41.7%) | 0.6 (0.2-65.1) | 0 | 12 (100.0) |
| **Subgroup 4b - Generalized myasthenia gravis** | 45/49 (91.8%) | 840.3 (233.6-2003.1) | 1 (2.0) | 42 (85.7) |
